# Characterization of the indoor near-field and far-field aerosol transmission in a model commercial office building

**DOI:** 10.1101/2021.02.25.21252239

**Authors:** Chih-Hsiang Chien, Mengdawn Cheng, Im Piljae, Kashif Nawaz, Brian Fricke, Anthony Armstrong

## Abstract

To simulate the exposure potential of infectious aerosol such as SARS-CoV-2 in an office building setting, experimental studies for airborne particle transmission have been conducted in a model commercial office building at the Oak Ridge National Laboratory. The synthetic test aerosol particles had diameters similar to that of viral particles, in the nanometer size range of genetic fragments. Thus, the test aerosol provided a realistic representation of SARS-CoV-2 viral particle transmission. The study results, which are still being analyzed carefully at the present, suggest that in a door-closed single room setting, the heating, ventilating, and air-conditioning (HVAC) system can facilitate aerosol transmission, and 10 measuring points in a single room report the normalized concentration ranged from 0.45 – 0.66. Additionally, at a measuring point 6 feet away from the source, the aerosol concentration can reach a plateau normalized concentration of about 0.6 within 30 minutes. When interior doors were closed, aerosol particle transmission into adjacent rooms occurred through the building HVAC system, at a lower rate compared to the open-door scenario. If the interior doors were open, however, then the transmission into adjacent rooms depends on building indoor air movement and distance from the source. The building HVAC system provided an approximately less than 10% aerosol transmission rate, while transmission through a door opening can add up to 40% of transmission into adjacent rooms from the source location.

**Footnote:** This manuscript has been authored by UT-Battelle, LLC, under contract DE-AC05-00OR22725 with the US Department of Energy (DOE). The US government retains and the publisher, by accepting the article for publication, acknowledges that the US government retains a nonexclusive, paid-up, irrevocable, worldwide license to publish or reproduce the published form of this manuscript, or allow others to do so, for US government purposes. DOE will provide public access to these results of federally sponsored research in accordance with the DOE Public Access Plan (http://energy.gov/downloads/doe-public-access-plan).

## Introduction

The nature of SARS-CoV-2 transmission suggests that in addition to close contact and droplets transmission, airborne transmission can contribute to human infection of this respiratory virus (CDC, 2021). The detection of SARS-CoV-2 RNA in aerosols (Santarpia et al., 2020) and the discovery of viable SARS-CoV-2 in air (Lednicky et al., 2020) support the airborne transmission route of the virus. Epidemiology studies have also reported several aerosol transmission events. Li et al. (2020) reported the aerosol transmission event of SARS-CoV-2 in a Guangzhou restaurant due to poor ventilation. The superspreading event at a choir rehearsal in Skagit Valley Chorale is likely due to inhalation of respiratory aerosol in the indoor environment (Miller et al., 2020). Airborne transmission of SARS-CoV-2 can be efficient under circumstances including prolonged exposure to respirable particles and inadequate ventilation or air handling (CDC, 2021), and these circumstances are relevant to indoor environments.

Due to the increasing demand to reduce energy consumption of buildings, the focus of heating, ventilating, and air-conditioning (HVAC) system design has been on energy efficiency and occupant comfort (Bhagat et al., 2020). The general strategy for building ventilation is to mix return air with fresh outside air (OA) to maintain indoor air quality. The ratio of return air to outside air can significantly impact HVAC system energy consumption and the indoor air quality. In winter, less OA is usually introduced into the building, because more energy will be required to heat the cold air mixture to achieve desired comfort. As the amount of recirculated air dominates the mixed indoor air, respiratory aerosols from occupants can recirculate and accumulate within the building, and the mitigation of aerosol transmission is subject to filtration in the air handling equipment. The ANSI/ASHRAE Standard 52.2-201 guideline indicates that Minimum Efficiency Reporting Value (MERV) 5-8 filters can provide appropriate air filtration for industrial workplaces. Unfortunately, MERV 5-8 filters have limited capacity to capture viral aerosols. For example, the efficiency of a MERV 8 filter for filtering 1 µm particles is approximately 30%, while the efficiency of a MERV 6 filter is about 18%. Therefore, there has been a rising concern about the health impacts of indoor aerosols in workplace buildings or other indoor environments with similar air handling and filtration equipment.

To slow SARS-CoV-2 spread, social distancing guidelines state that individuals must stay at least 6 feet (about 2 meters) from others who are not from the same household in both indoor and outdoor spaces (CDC, 2021). The effectiveness of the 6-foot guideline to slow respiratory aerosol spread is still under debate. The 6-foot guideline was based on an outdated dichotomous concept of respiratory droplet size (Jones et al., 2020), while droplets of all sizes are moved by the exhalation. The turbulent cloud from natural human forceful emissions such as a sneeze can reach over 8 meters (Bourouiba, 2020), which is greatly beyond the 6-foot guideline.

In indoor zones with mixing ventilation, OA and return air are mixed to provide thermal comfort. The mixing, however, can facilitate the transmission of aerosols in that zone; thus, the extent of near-field aerosol transmission must be quantified. Second, because supply air largely consists of recirculated air, in a building with multiple zones, it is possible that aerosols from the source zone can be transmitted to other zones through the centralized HVAC system. The degree of such a far-field aerosol transmission has not yet been reported.

To investigate these knowledge gaps, the specific goals of this study are to 1) experimentally characterize the 3-dimensional aerosol concentration distribution in the source zone and 2) experimentally characterize the cross-room aerosol transmission under varying indoor air environments.

This article is being released before peer review, recognizing the need for timeliness of this information to the public for risk evaluation.

## Methodology

In this study, sodium chloride aerosols were chosen as the preferred surrogate for respiratory aerosols because of the safety concerns of using bioaerosols during indoor air experiments. Sodium chloride aerosols tagged with Uranine (fluorescent marker) were generated and collected in cascade impactors to map and quantify the near-field and far-field aerosol transmission under the influence of a centralized Variable Air Volume (VAV) HVAC system in a model commercial office building.

### Building and Indoor Air Ventilation

The aerosol transmission experiments were conducted in a model commercial office building, the Flexible Research Platform (FRP), at the Oak Ridge National Laboratory during October and November 2020. The FRP was designed to represent a typical two-story small-to-medium office building that was built in the 1990s. The two-story FRP has a total floor area of 3,200 ft^2^ for investigating building energy efficiency and has been equipped with more than 500 sensors and instruments to monitor the building environment, such as relative humidity (RH) and temperature and HVAC system and building performance. This test platform has been used for multiple studies, including empirical validation of the building model and HVAC system performance analysis (Im et al., 2020; Lee, Im, and Song, 2018). Hence, the building’s envelope and HVAC system properties, such as the building’s air infiltration rate and HVAC system performance, have been well characterized, which can be leveraged in this multizone aerosol transmission study. In addition, the building has emulated occupancy using humidifiers and ceramic fan-forced heaters.

The total volume ventilation strategy in this study is to use mixing ventilation. Since MERV 7 and 8 filters are commonly used in centralized HVAC systems in office buildings, a VAV centralized HVAC system with such filters (Figure 1) was used to investigate the effect of the centralized ventilation system on aerosol transmission. Additionally, several measuring points for humidity, temperature, and air flow rate were available to help understand the indoor aerosol transmission.

**Figure 1.**
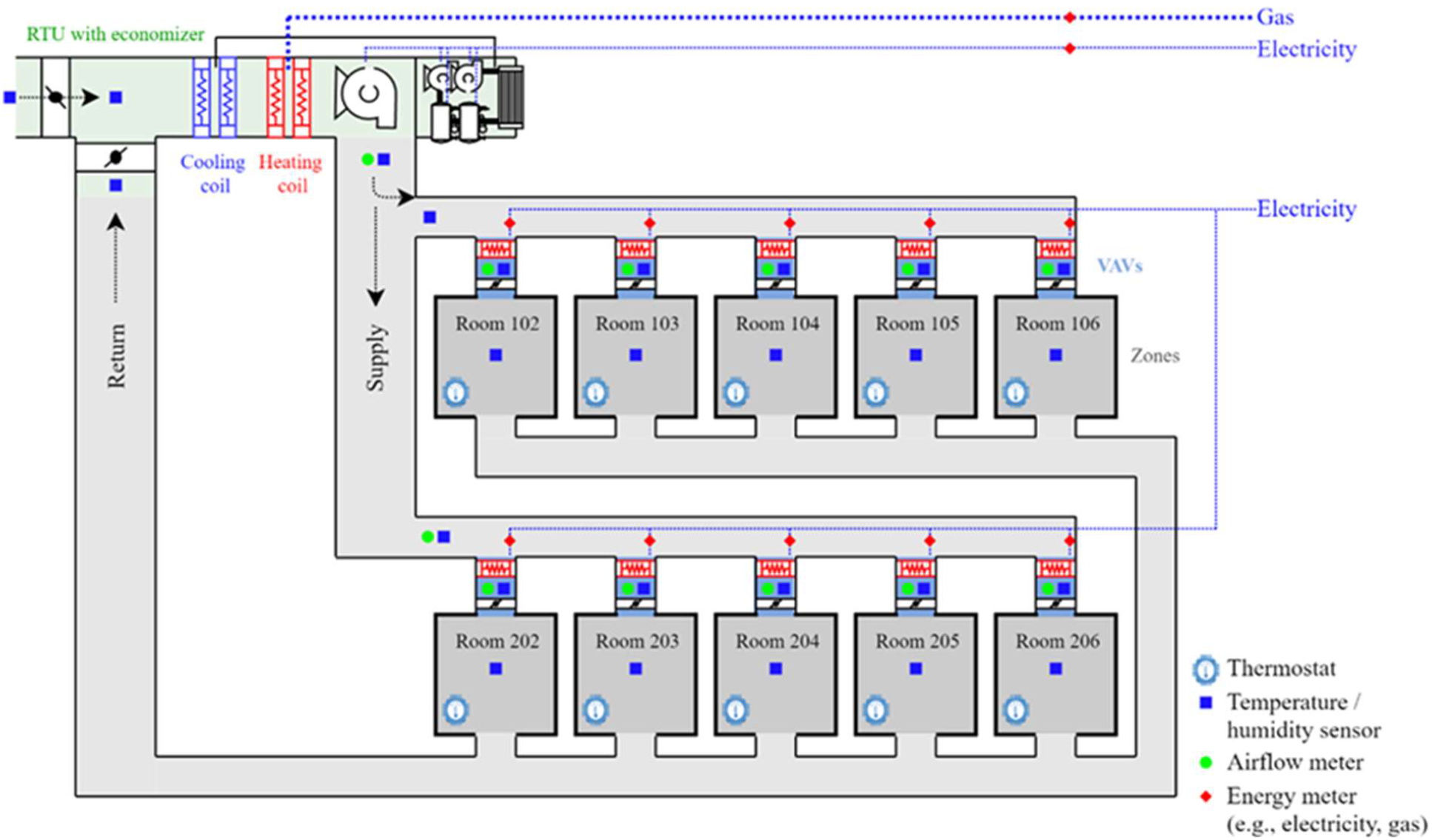
Variable air volume (VAV) centralized HVAC system configuration and measuring points

Figure 2 shows the first floor, composed of five zones (one core zone and four surrounding perimeter zones) with return, supply, and exhaust air vents. The exhaust fan extracts air from the core zone directly and not from the core plenum. The air from each zone enters the plenum space of each room through the return grille and then travels to the return duct through the plenum. OA mixes with the return air; after filtration, the mixed air is conditioned through a cooling and/or heating coil and then directly dispersed into each zone via the supply duct system. The thermostat of the HVAC system was set to maintain the temperature between 70 to 76°F. The minimum and maximum supply air flow rate for each zone varies per size and orientation of the room, and the VAV box in each room modulates the air flow rate to meet the cooling demand of the room. The air flow rate for the five rooms in the first floor ranges from 120 to 600 CFM. In general, air exchanges per hour (ACH) ranges from 5 – 8.

**Figure 2.**
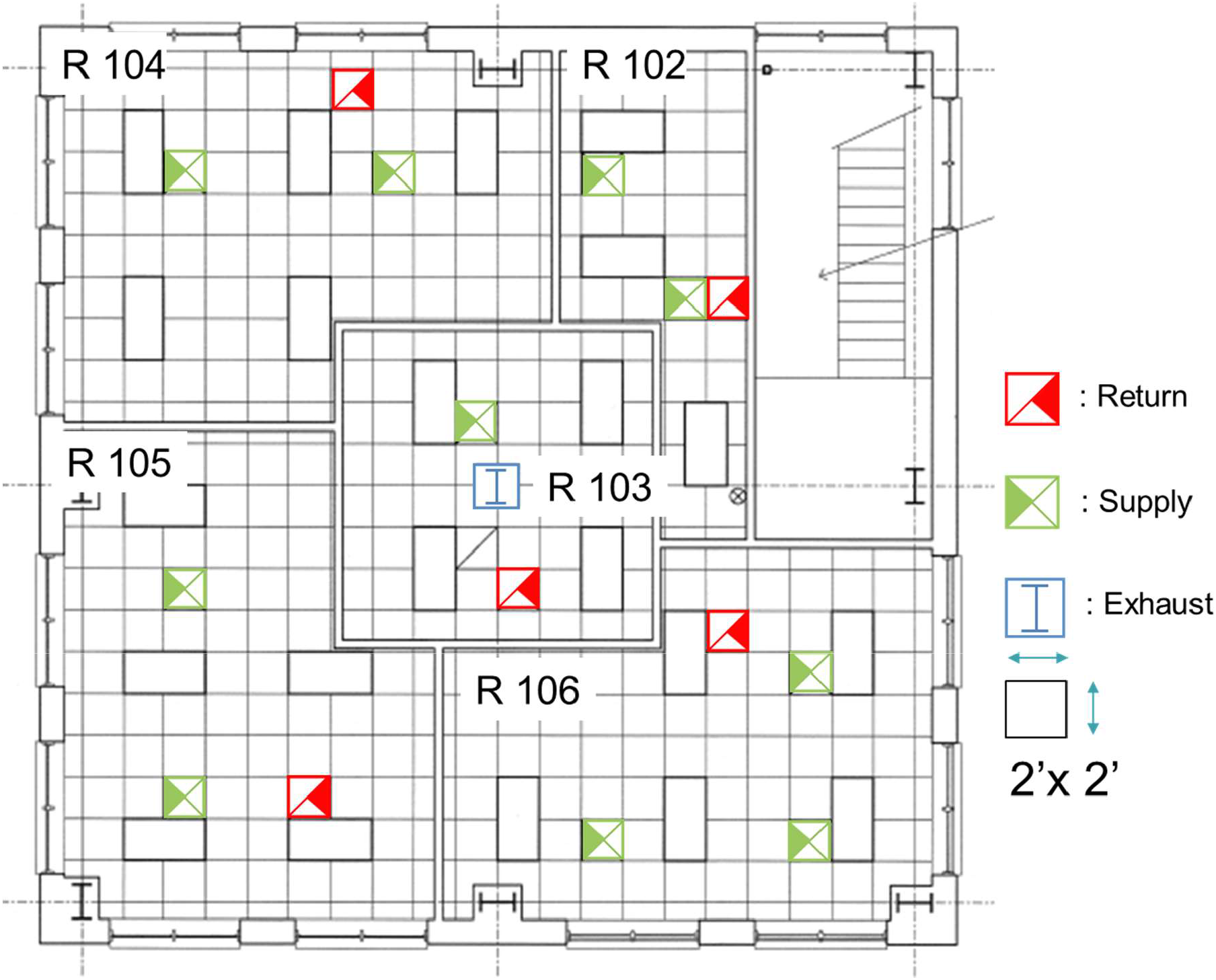
Locations of return, supply, and exhaust air on the first floor in the FRP

### Aerosol generation, collection, and measurement

Sodium Chloride (MW = 58.44 g mol^-1^; NaCl; CAS#: 7647-14-5; Sigma Aldrich) was selected as the material for testing aerosol transmission in this study. As the NaCl droplets initially leave the aerosol generator, the NaCl droplets evaporate and turn into solid aerosols in a short time. The size of these solid chloride aerosols does not change, because the relative humidity (RH) in the building environment has been controlled to be lower than the deliquescence RH (75%). To positively identify generated aerosols against the background indoor aerosols or aerosols from the OA with the same size, Uranine (Fluorescein sodium salt; MW= 376.27 g mol^-1^; C_20_H_10_Na_2_O_5_; CAS#: 518-47-8; Sigma Aldrich) was tagged into the sodium chloride aerosol to provide a unique fluorescent signal for aerosol characterization from the ambient environmental aerosols. Similar tagging technology has been used to understand indoor aerosol transmission in aircraft (Kinahan et al., 2021). The nebulization solution was prepared by dissolving 3 g NaCl and 1 g Uranine into 400 mL Nanopure water. The TSI model 3074 was used for the particle generation, because it can generate sufficient aerosols with constant output (Liu and Lee, 1975). During the experiments, the nebulizer continuously generated aerosol during the sampling period.

Sioutas cascade impactors were used to collect fluorescent aerosol, because high flow rate at 9 Lpm can collect sufficient sample for mass analysis within a reasonable time frame. The cascade impactors classify aerosols into five stages based on their aerodynamic diameter, ranging as follows: <0.25 µm, 0.25 – 0.5 µm, 0.5 – 1.0 µm, 1.0 – 2.5 µm, and > 2.5 µm (Misra et al., 2002). Particles are collected on Polytetrafluoroethylene (PTFE) (25-mm, 0.5 µm pore size) substrates and a final PTFE filter (37-mm, 2.0 µm pore size). The sampling time depends on source concentration, room dimension, ventilation condition, and analytical quantification limit. Several experiments concluded that at least one hour of sampling time was required to collect adequate particle mass for analysis.

Particles on filters were extracted after each campaign and dissolved in a cuvette with Nanopure water for fluorescent analysis. Water is a good extracting solution for aerosol tagged with Uranine on filters, as the extraction efficiency is > 99% (Tolocka, Tseng, and Wiener, 2001). The fluorescent signal of aerosol was detected by a custom-made fiber-optics coupled spectrometer with an LED excitation wavelength of 470 nm, and the fluorescent emission was detected at 525 nm with a high-resolution spectrometer (OceanInsight Model HR4000CG-UV-NIR). The spectral signal was recorded and processed by the OceanView software on a 64-bit Windows-based laptop. The quantification limit by this spectral system was determined, prior to the campaigns, to be approximately 0.3 µg, which was sufficient for the intended fieldworks.

In addition to the impact of HVAC, the different particle releasing rates from the aerosol generator can contribute to variation of absolute aerosol concentration. To normalize the concentration variation, a normalized concentration (NC) was defined as follows:

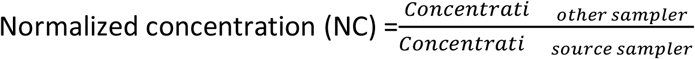

NC serves as an indicator to evaluate the relative exposure level at any location in reference to the source. NC ranges from 0 to 1, and a value of 1 indicates that the concentration at a particular location is the same as the source concentration.

### Experimental design: near-field and far-field campaign

Experiments were conducted during October through November 2020. Fluorescent particles in the submicron range were generated and monitored at various locations throughout the campaign. Near-field and far-field experiments were undertaken in this study. Each experiment was conducted on different days, with sufficient time between experiments to minimize the influence of one test on the next.

For the near-field studies, the goal was to quantify the indoor aerosol transmission in a single room under the influence of a VAV centralized HVAC system with a mixing ventilation strategy. Three monitoring towers were placed 6 ft, 8 ft, and 10 ft away from the aerosol source, and each tower was equipped with three cascade impactors at heights of 3 ft, 5 ft, and 6 ft above the floor. The 4^th^ tower was 1 ft away from the aerosol source and equipped with the 10^th^ cascade impactor at a height of 3 ft above the floor (Figure 3). The 10^th^ cascade impactor sample represents the source concentration, while the other nine cascade impactor samples were used to characterize the three-dimensional distribution of aerosol mass concentration in the room. Real-time aerosol measurement instruments, such as a Scanning Mobility Particle Sizer Spectrometer (SMPS, TSI Model 3080L + 3025A), an Aerodynamic Particle Sizer (APS, TSI Model 3320), and a AeroTrak (TSI Model 9110-01), were deployed in this zone to monitor the aerosol concentration.

**Figure 3.**
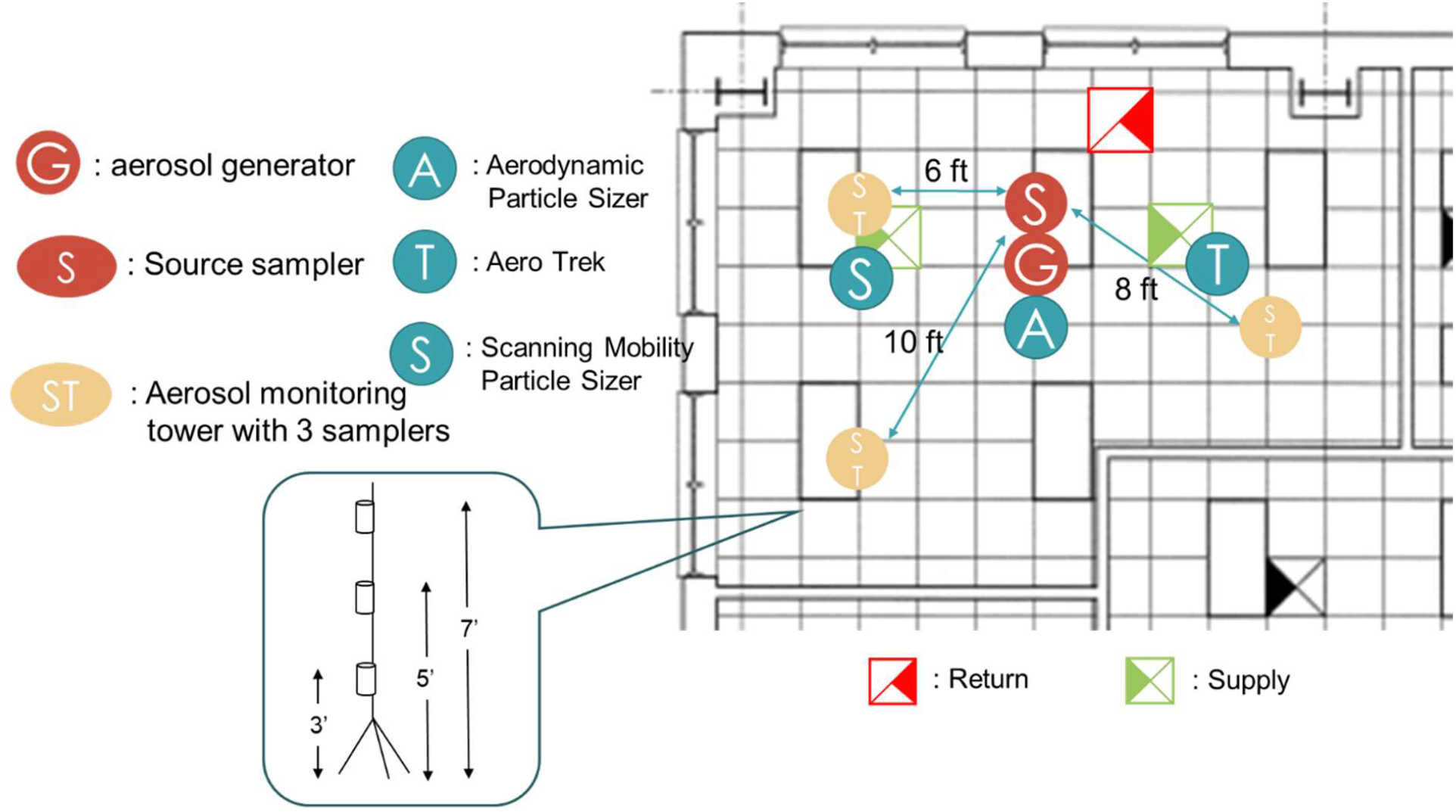
Location of samplers in the near-field campaign

The far-field campaigns were designed to quantify the aerosol transmission from the source room (R104, Figure 4) to the other rooms on the first floor under the influence of a centralized HVAC system. Open- and closed-door scenarios were tested. Closed-door scenarios represent an office setting in which aerosols are mainly transported by the HVAC system and possibly through door gaps; in the open-door scenario, door-gap transmission is maximized. The location of the aerosol generator and samples for both scenarios are shown in Figure 4. The aerosols were generated in the source room (R104), and a source impactor was 1 ft horizontally away from the aerosol generator and at a height of 3 ft above the floor. Impactors 2, 3, 4, and 5 were located in rooms R103, R102, R105, and R106; these impactors were deployed at a height of 3 ft above the floor and 5 ft away from the door. As supply air can dilute aerosol concentrations in the vertical direction, as shown in the near-field study, impactors 6, 7, 8, 9, and 10 were at a height of 7 ft and located under the four-way square ceiling diffusers to sample aerosol concentration under VAV diffusers in rooms R104, R103, R102, R105, and R106, respectively. Again, several real-time instruments were deployed in the source room to monitor the aerosol concentrations for Quality assurance (QA) and quality control (QC) purposes.

**Figure 4.**
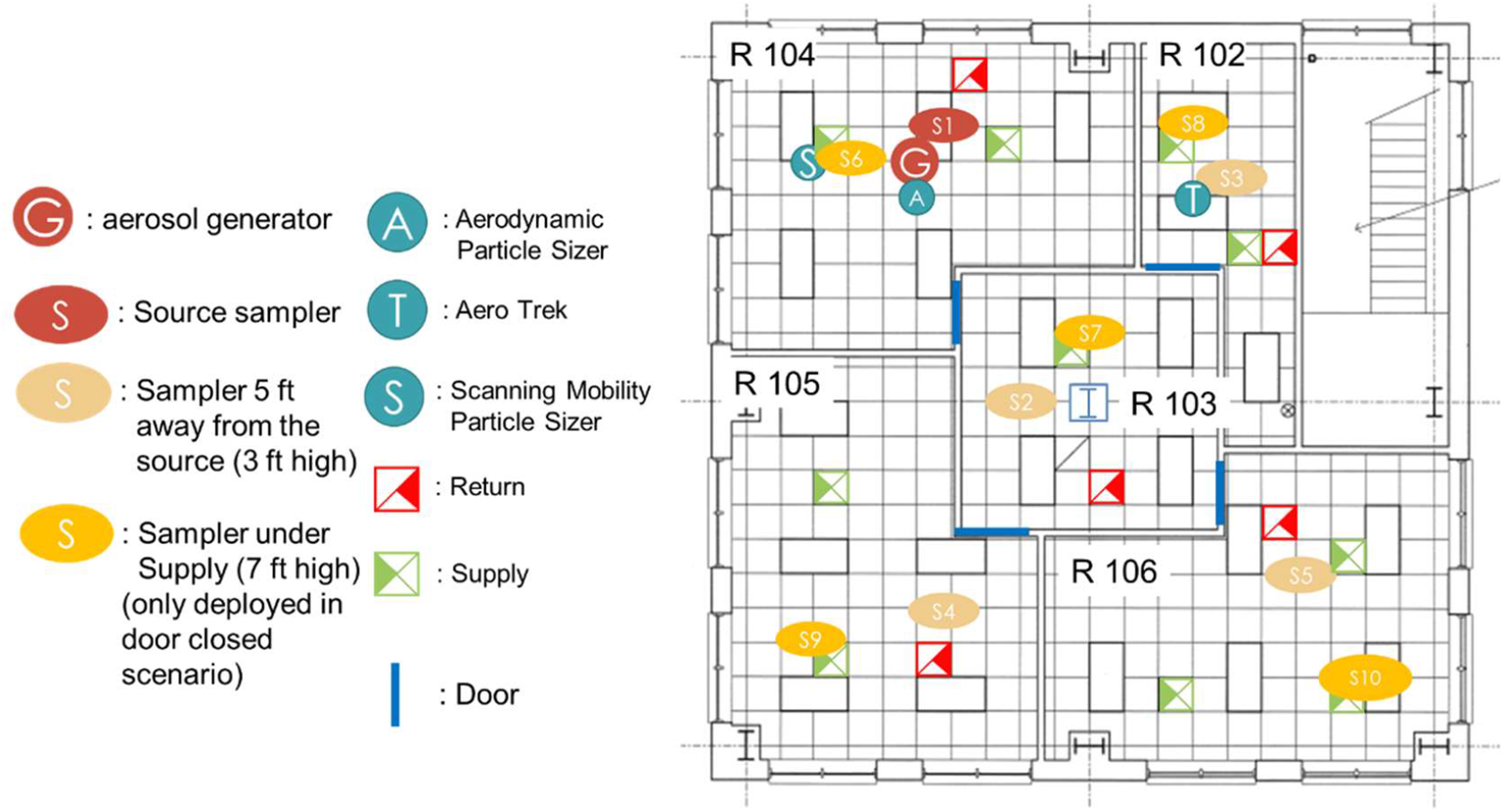
Location of samplers in the far-field campaign

## Results and discussion

### Near-field campaigns

The near-filed campaign was conducted from 10 am to 2 pm on October 27, 2020. The relevant building environment, given in Table 1, indicates the room temperature and RH were 73±1 °F and 48±2 %, respectively, during the sampling period. Note that the air exchange rate in Table 1 refers to the recirculated air. The average VAV flow rate in R104 is 321 CFM, which is equivalent to the air exchange rate of 7.7 (h^-1^).

**Table 1.** Building environment for the near-field campaign.

| Space | R104 (source) | Outdoor |
| --- | --- | --- |
| Room Temperature (°F) | $73 \pm 1$ | $64 \pm 3$ |
| RH (%) | $48 \pm 2$ | $81 \pm 4$ |
| VAV flowrate (CFM) | $321 \pm 2$ | NA |
| Volume (ft<sup>3</sup>) | 2496 | NA |
| Air exchange per Hour | 7.7 | NA |

Real-time submicron measurement using SMPS demonstrated that the aerosol generator can maintain constant output for four hours (Figure 5). Fifteen minutes after the start of aerosol generation, the aerosol concentration at the 6 ft distance from the source reached a steady state value, with aerosol size ranges from 100 nm to 500 nm.

**Figure 5.**
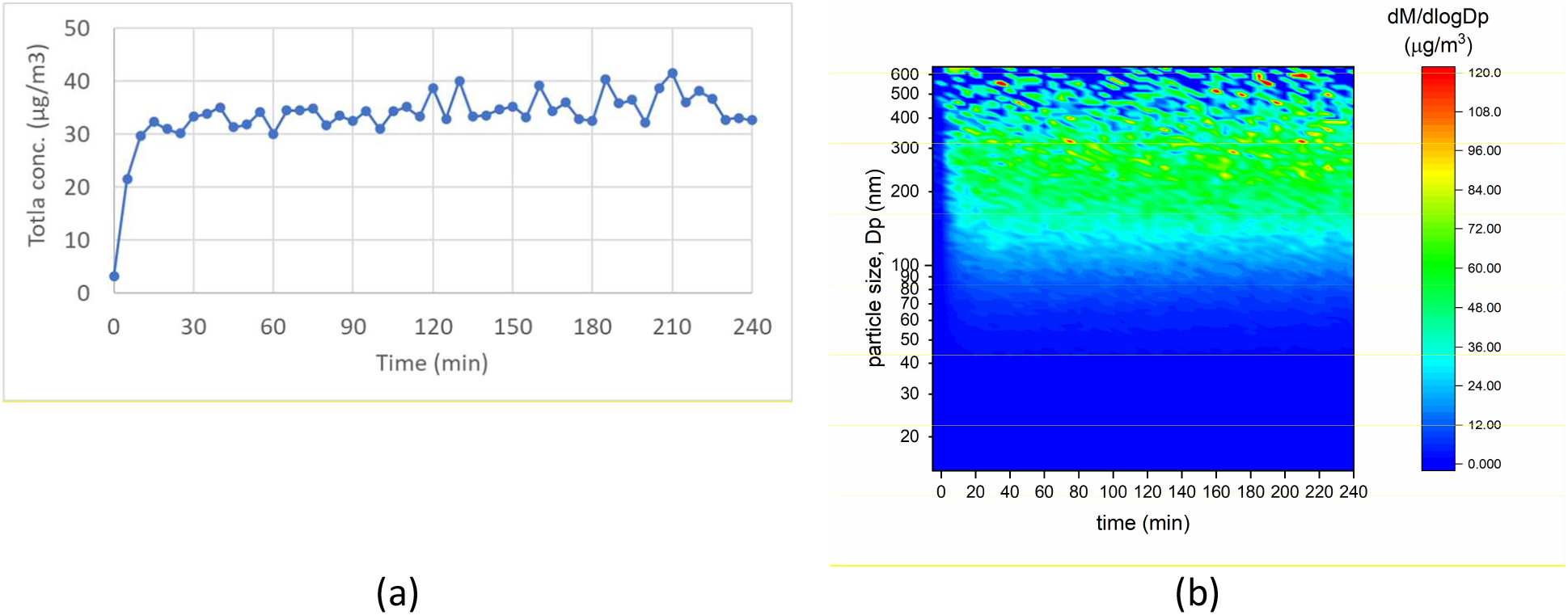
The total mass concentration (a) and size distribution over time (b) in the source room measuring sodium chloride testing aerosol using TSI model 3074 aerosol generator

The concentration distribution measured using the 10 impactors is shown in Figure 6. Figure 6(a) indicates the total mass source concentration is approximately 80 µg/m^3^; as expected, the size distribution is dominated by submicron aerosols. The other impactors collected fewer particles, but submicron particles still dominated the size distribution (Figures 6[b]-[d]). Figure 7 displays the normalized concentrations distributed in the source room. Only submicron fraction was calculated. Figure 7 demonstrates the NC is as high as approximately 0.6, regardless of the distance of the cascade impactors from the source. In addition, Figure 5 shows the plateau concentration is reached within 30 minutes. Therefore, even if occupants are in compliance with the 6-foot rule or even when occupants are spatially greater than 6 feet apart, the exposure risk is still high when occupants share the same zone with an index patient.

**Figure 6.**
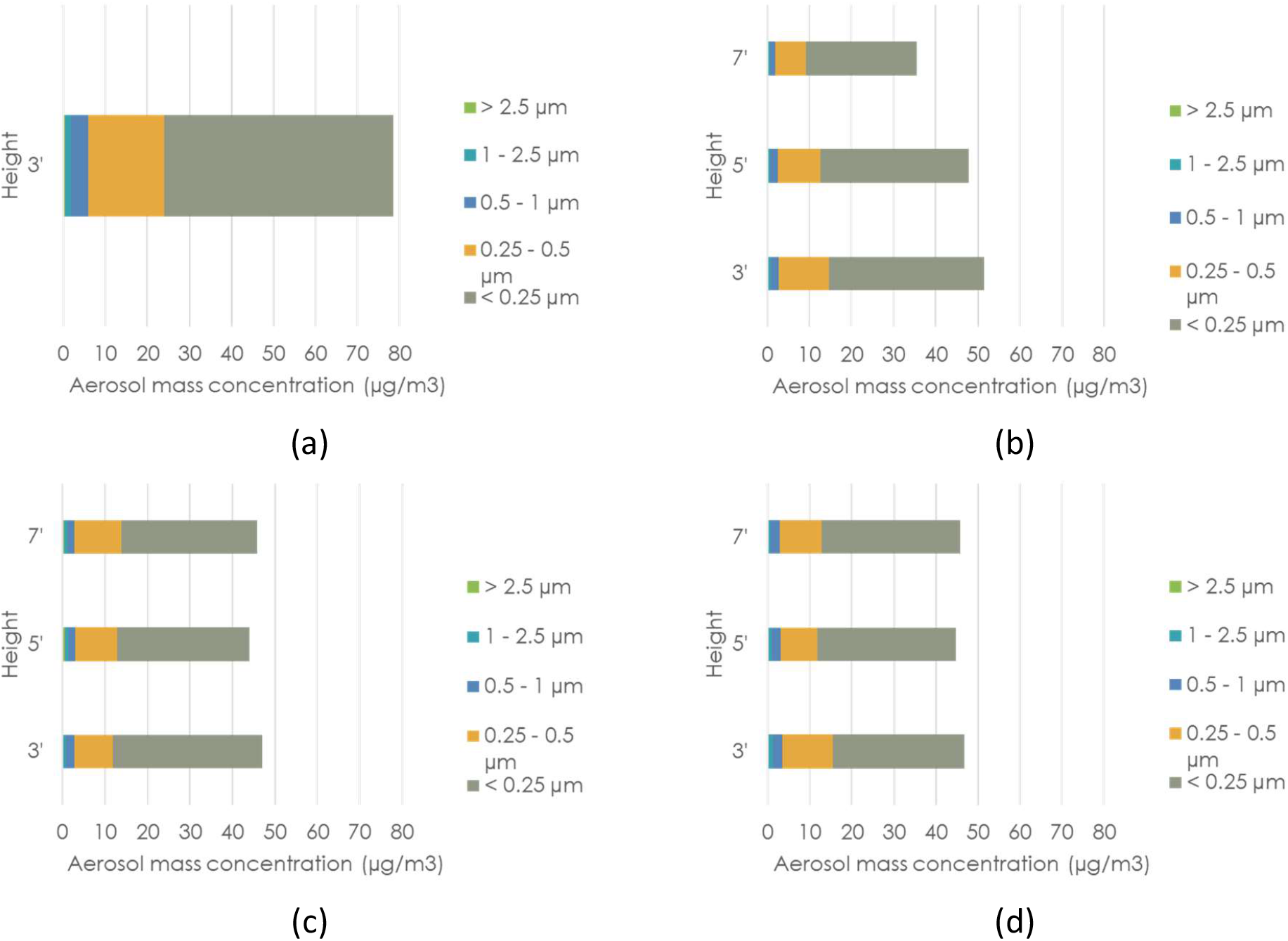
The spatial distribution of aerosol concentration measured at (a) source, (b) 6 ft, (c) 8 ft, and (d) 10 ft away from the source in the horizontal direction.

**Figure 7.**
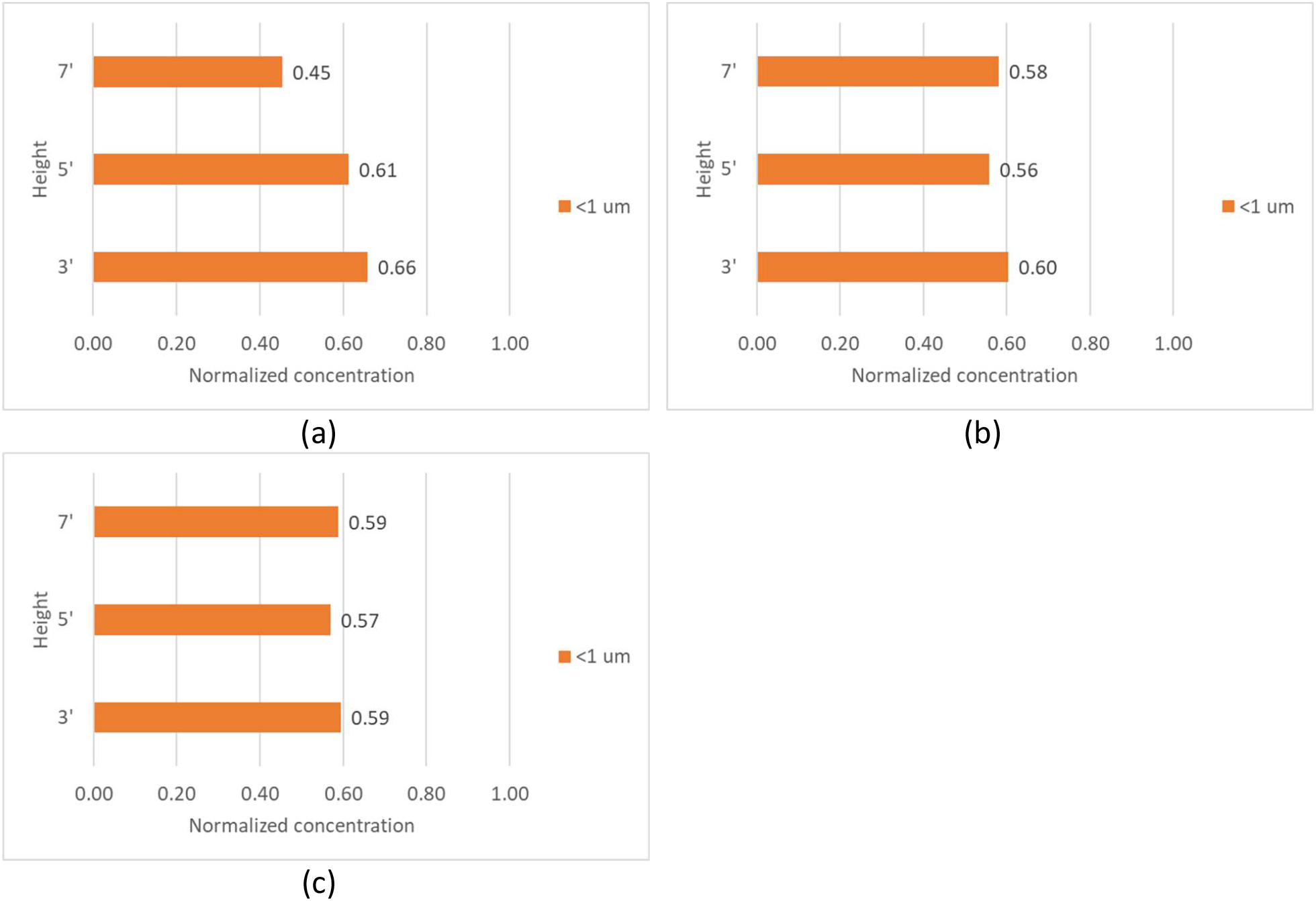
The spatial distribution of normalized concentration measured at (a) 6 ft, (b) 8 ft, and c) 10 ft away from the source in the horizontal direction.

The coefficient of variation (CV), the ratio of standard deviation to the average of sample concentrations, was used as an indicator to quantify the concentration variation in the vertical direction and on the horizontal plane. For samples in the sampling towers 6 ft, 8 ft, and 10 ft away horizontally from the source, the coefficient of variation (CV) is 19%, 4%, and 2%, respectively. On the other hand, CV on the horizontal plane at 3 ft, 5 ft, and 7 ft height is 6%, 5%, and 14%, respectively. The higher CV (i.e., 19% and 14%) is attributed to the lower concentration measured at the 7 ft height and 6 ft away from the source. When the impactors are located below the diffusers, the aerosol concentration decreases as the height increases. Although the supply air is a mixture of recirculated air and OA and the filtration effect is limited, the aerosol concentration in the supply vent air is still lower than the indoor environment. This less-contaminated supply air dilutes the aerosol concentration. On the other hand, at 10 ft and 8 ft away from the source in the horizontal direction, the data are less susceptible to the jet flow from the diffuser. Additionally, the ceramic fan-forced heaters that were set to emulate human occupancy may provide local air convention and buoyancy forces associated with temperature difference, enhancing the local mixing of aerosols.

### Far-field campaigns

The open-door far-field campaign was conducted from 9 am to 1 pm on November 3, 2020, and a closed-door campaign was performed from 9:15 am to 1:15 pm on November 10, 2020. Indoor conditions such as room temperature, VAV flowrate, and ACH, shown in Table 2, are similar for the two experiments. Room RH in the closed-door experiment ranged from 20% to 23%, while RH in the open-door scenario ranged 42% to 51%. As the OA temperature during the open-door experiment was lower than the closed-door experiment, the reheat coil in the HVAC system warmed the supply air, resulting in a lower RH measured in the zones by approximately 21%. The VAV supply flow rates vary in zones, as shown in Table 2, and the lowest flow rate occurs in R102. The supply air ACH in the zones also varies, but the lowest ACH is not in R102 because ACH is determined by not only the flowrate but also the zone volume. The lowest supply air ACH of 5.3 occurred in the core zone. Additionally, the exhaust fan (Figure 2) directly extracted air from the core zone at approximately 250 CFM.

**Table 2.**
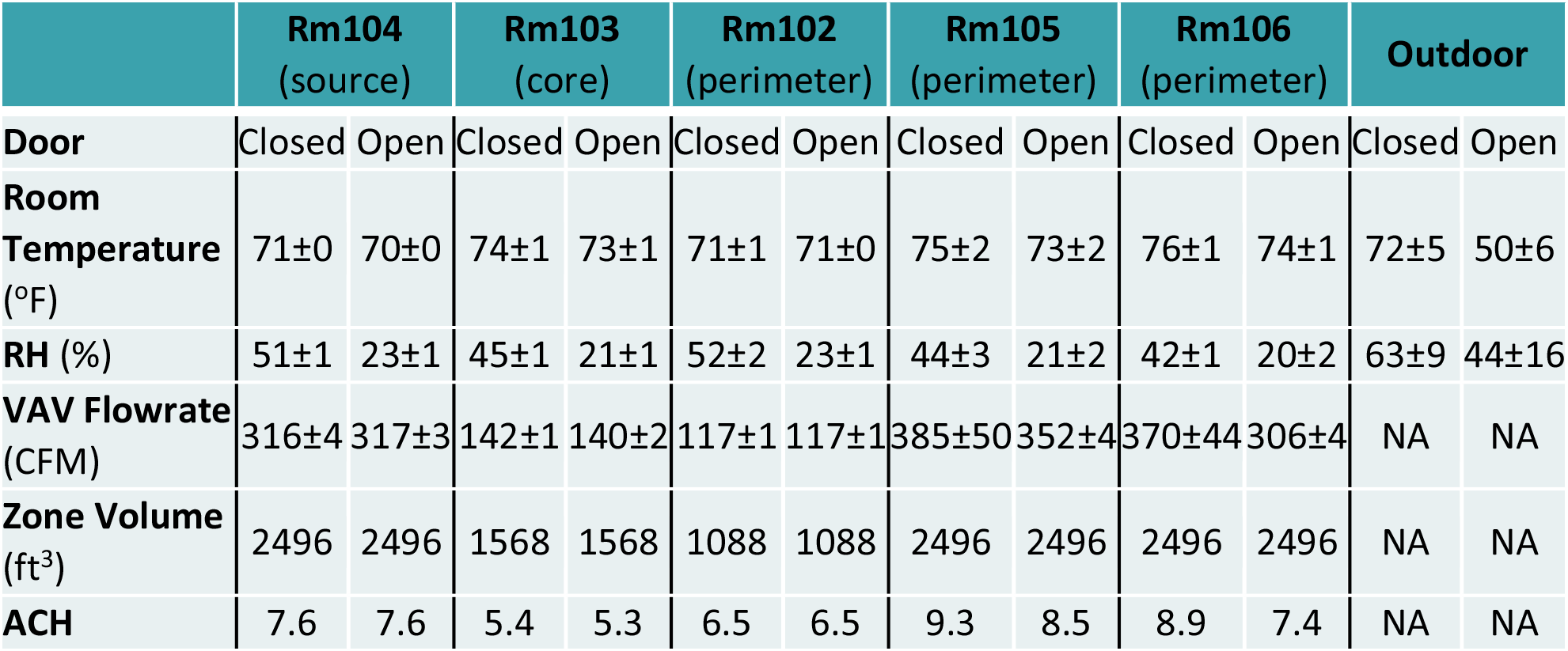
Building environment for the far-field campaign

Figure 8 displays the aerosol concentrations distributed in the five zones for the open-door and closed-door scenarios. The total mass concentrations from the source were approximately 65 µg/m^3^ for the two scenarios, and submicron aerosols still dominated mass concentration.

**Figure 8.**
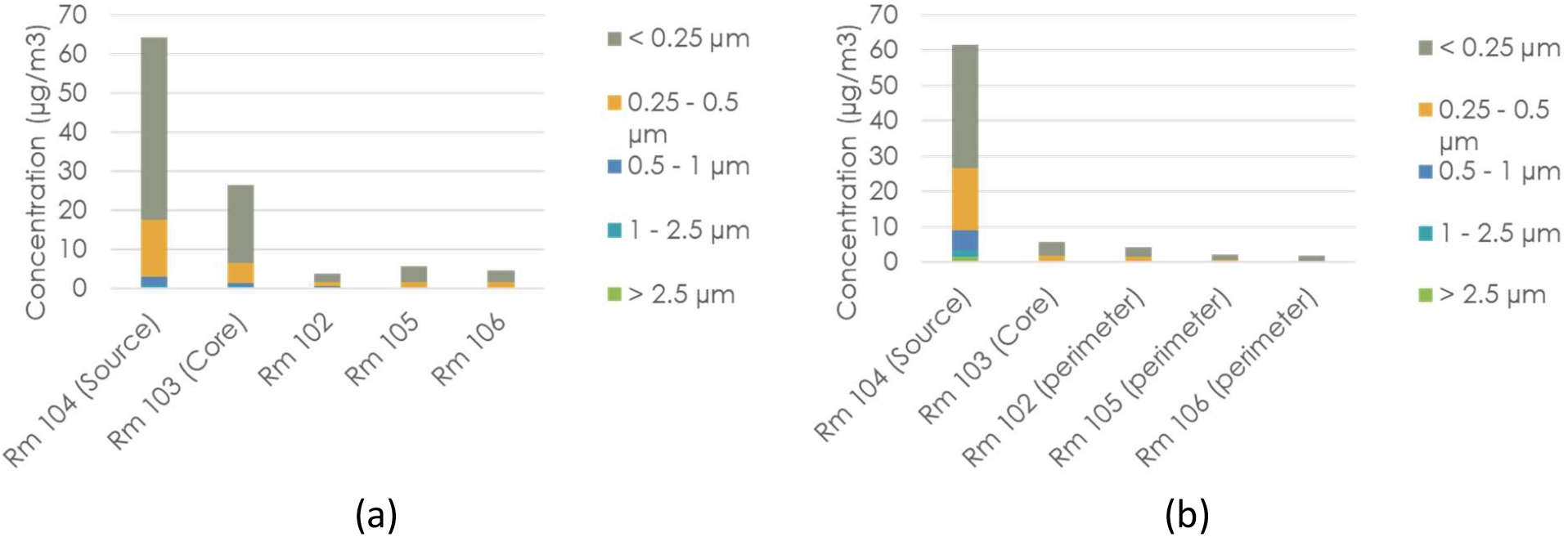
The spatial distribution of aerosol concentration for (a) open-door and (b) closed-door scenarios

Figure 9 shows the spatial distribution of normalized concentrations for the submicron particles. The results show the normalized concentration in the core zone is higher than the perimeter zone. As the exhaust fan draws more air than the supply from the core zone (R103), negative pressure draws aerosols from the perimeter zones through the door opening to the core zone. The exposure risk in the core zone can be reduced from 41% to 10% if the doors were closed. In addition, the normalized concentration values in the perimeter zones were less than 10%, irrespective of door-opening condition.

**Figure 9.**
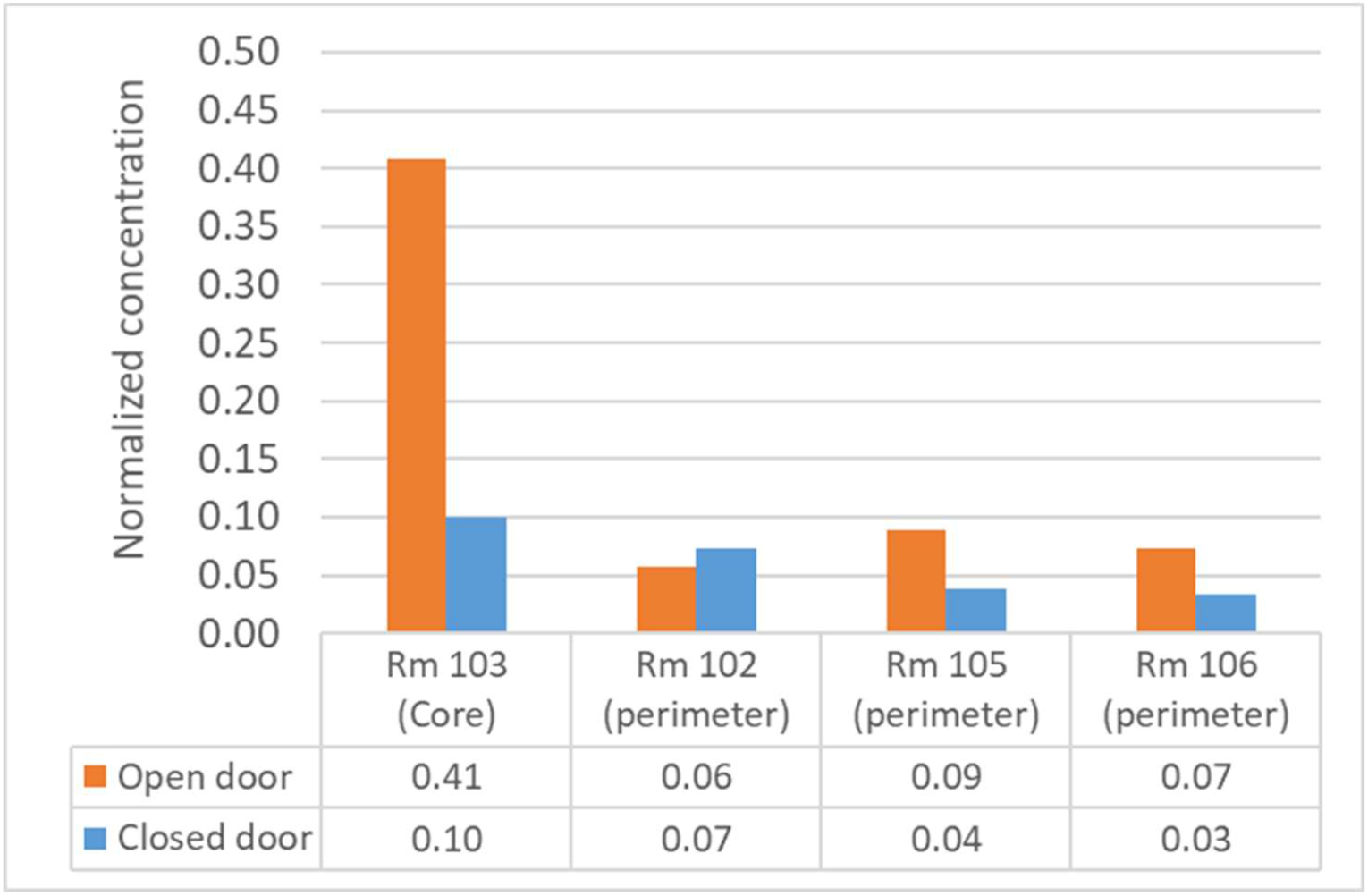
The spatial distribution of submicron normalized concentration

Figure 10 reports the aerosol concentrations under the diffusers in the five zones (Impactor 6 – 10 in Figure 4). Total mass aerosol concentrations under the diffusers in Rooms 103 and 102 are 3 µg/m^3^, while the concentration under the diffusers in Rooms 105 and 106 are 2 µg/m^3^. The concentration difference in these non-source rooms are insignificant, indicating the aerosol concentration in the supply air is about 2.5 µg/m^3^. However, the aerosol concentration under the diffuser in the source room is 17 µg/m^3^, much higher than the concentrations in non-source rooms. This implies the aerosol concentration under the diffuser in the source room is from both the supply air and the aerosol generator. In other words, fresh particles emitted from the aerosol generator entrained and mixed with supply air in the sampling zone under the diffuser. As the source concentration of 62 µg/m^3^ and the aerosol concentration in the supply air of 2.5 µg/m^3^ were measured, the aerosol transmission through the HVAC is concluded to be 4%.

**Figure 10.**
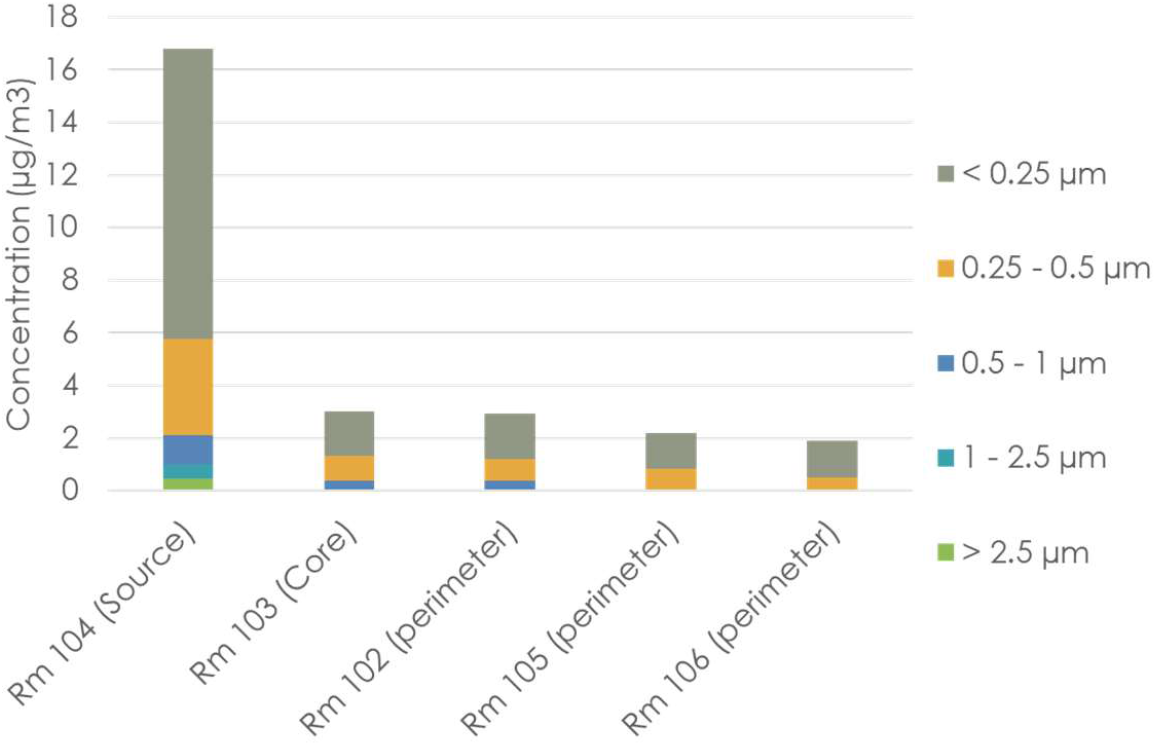
Aerosol concentrations under diffuser in the closed-door scenario

## Data Availability

All relevant data are within the manuscript.

## Acknowledgements

This research was supported by the Department of Energy/Office of Energy Efficiency and Renewable Energy Research. Anthony Gehl and Seungjae lee are acknowledged for assistance in experimental setup and building sensors. This research used resources at the Building Technologies Research and Integration Center, a DOE Office of Science User Facility operated by the Oak Ridge National Laboratory. Oak Ridge National Laboratory is managed by UT-BATTELLE, LLC for the U.S. Department of Energy under contract DE-AC05-00OR22725.

